# Systematic review of cardiac adverse effects in children and young people under 18 years of age after SARS-CoV-2 vaccination

**DOI:** 10.1101/2021.12.06.21267339

**Authors:** Joana Cruz, Amedine Duret, Rachel Harwood, Lorna K. Fraser, Caroline B. Jones, Joseph Ward, Elizabeth Whittaker, Simon E. Kenny, Russell M. Viner

## Abstract

**Background:** Reports of myocarditis and pericarditis following vaccination with mRNA vaccines for SARS-CoV-2 have occurred after countries began vaccinating adolescents. We undertook a systematic review of cardiac adverse effects associated with SARS-CoV-2 vaccine in children and young people (CYP)< 18 years.

**Methods:** Systematic review with protocol prospectively registered with PROSPERO (CRD42021275380).

Six electronic databases were searched from 1 December 2019 to 14 September 2021. Eligible studies were those reporting on CYP with reported or proven myocarditis, pericarditis and/or myopericarditis associated with vaccination against SARS-CoV-2. We summarized findings across all clinical cases reported in case report / case series studies. As a number of studies reported data from two publicly available vaccine surveillance systems, we updated estimates of reporting rates for cardiac adverse events up to 31 October for the US Vaccine Adverse Event Reporting System (VAERS) and 13 November for EudraVigilance covering European Union and European Economic Area (EUEA) countries.

**Results:** Twenty-one studies were included from 338 identified records. Seventeen were case reports/series describing a total of 127 CYP. Three studies described reporting rates from passive surveillance databases (VAERS, EudraVigilance, and the WHO VigiBase) and one described 22 cases from the US Vaccine Safety Datalink (VSD).

Clinical series reported that 99.2% presented with chest pain, 100% had raised troponin and 73.8% had an abnormal ECG. Cardiovascular magnetic resonance (CMR) in 91 cases identified myocardial injury in 61.5%, with 90.1% showing late gadolinium enhancement. NSAIDs were the most common treatment (76.0%).

One US dataset (VSD) estimated a significant excess of 29.6 events per million vaccine doses across both sexes and doses. There were 1129 reports of myocarditis and 358 reports of pericarditis from across the USA and EU/EEA. The VAERS reporting rate per million for myocarditis was 12.4 for boys and 1.4 for girls after the first dose, and 49.6 for boys and 6.1 for girls after the second dose. There was a marked trend for VAERS reporting to be highest soon after initiation of the vaccine schedule, suggesting reporting bias.

**Conclusions:** Cardiac adverse effects are very rare after mRNA vaccination for COVID-19 in CYP <18 years. The great majority of cases are mild and self-limiting without significant treatment. No data are yet available on children under 12 years. Larger detailed longitudinal studies are urgently needed from active surveillance sources.

## Introduction

The introduction of vaccines to protect against serious illness in SARS-Cov-2 has been highly effective ^1^. However, there have been a number of case reports describing cardiac adverse effects, namely myocarditis, pericarditis and myopericarditis after SARS-CoV-2 vaccination ^2^ ^3^. From May 2021 onward, several countries extended vaccination to include children and young people (CYP) aged 12-15 years old ^4^ ^5^. Soon after this, reports were published of cases of myocarditis in CYP after SARS-CoV-2 vaccination, especially in males ^6^. Myocarditis and associated conditions have also been reported after SARS-CoV-2 infection, both with COVID-19 disease ^7-9^ and with the post-infectious paediatric multisystem inflammatory syndrome (PIMS) ^10^, also known as multisystem inflammatory syndrome in children (MIS-C).

By October 2021, several reports had been published with wide variation in reported frequency of myocarditis after vaccination ^9 11 12^. The frequency of myocarditis in these studies varied according to gender, age group and whether first or second dose administrated ^13^ ^14^.

Estimates across broader age-groups using high-quality data sources suggest the risk of myocarditis was highest amongst young men, with estimated incidence after the first dose of 106.9 per million amongst males 16-29 years,^11^ and 13.4 and 150.7 per million after first and second doses respectively.^12^ Males aged 12-15 years old appeared most affected, particularly after the second dose.^13^ ^14^

As of early November 2021, over 50 million doses of mRNA vaccines have been given to 12-17 year olds across the USA^15^ and European Union and European Economic Area^16^ countries, with an additional 2.8 million doses given in the UK^17^ and 3.1 million in Australia.^18^

Establishing the nature and frequency of vaccination adverse events such as myocarditis is essential to inform both national vaccination strategies and clinical decision-making for individuals, particularly as nations expand adolescent vaccination and consider vaccinating younger children. We therefore undertook a systematic review of cardiac adverse effects associated with SARS-CoV-2 vaccine in those under 18 years.

## Methods

We followed a standardised methodology and reported results according to Preferred Reporting Items for Systematic Reviews and Meta-Analyses (PRISMA) ^19^ recommendations. The protocol was prospectively registered with PROSPERO (CRD42021275380).

### Review question

What are the characteristics, frequency and natural history of cardiac adverse effects in children and young people (CYP) (<18 years old) after SARS-CoV-2 immunisation?

### Search strategy

We systematically searched PubMed, MedRxiv, Europe PMC, World Health Organisation’s COVID-19 Global Literature on Coronavirus Disease, Research Square and Google Scholar from 1 December 2019 to 14 September 2021 for peer-reviewed papers and reports on cardiac adverse events caused by SARS-CoV-2 immunisation in CYP. The search strategy used the keywords “myocarditis” OR “pericarditis” OR “adverse effects” AND “covid-19” AND “vaccin*” AND “children” OR “adolescent” OR “paediatric” in title or abstract of the reference. The complete list of key word strings for each database can be found in supplementary appendix (Table S1). The first 40 results from Google Scholar were also analysed, along with the databases of the US Vaccine Adverse Event Reporting System (VAERS)^20^ and Eudra-Vigilance—the European database of suspected adverse drug reaction reports ^21^ for cases of myocarditis, pericarditis or myopericarditis related with SARS-CoV-2 vaccine in CYP aged between 12-17 years old. We included adverse effects from mRNA-1273 (Moderna) and BNT 162b2 (Pfizer) vaccines since they were the only vaccines approved for the age group <18 years old at the time of writing of this paper.

### Eligibility

We included papers that reported myocarditis, pericarditis and/or myopericarditis associated with vaccination against SARS-CoV-2 in CYP, aged <18 years old. Studies that also included individuals older than 18 years old were included if they allowed extraction of data on eligible individuals and/or age groups separately. We excluded studies on CYP SARS-CoV-2 vaccination trials (being underpowered), systematic reviews that mentioned cardiovascular involvement but did not specify the above conditions, studies that included patients with cardiovascular conditions diagnosed prior to SARS-CoV-2 vaccination or that used non-human models. Where multiple studies reported data from vaccine surveillance databases, we included only the most recent studies to avoid duplication.

### Study selection

The PRISMA flowchart is shown in figure 1. Our search identified 330 articles to which we added eight articles identified through the snowball technique. Titles and abstracts of potentially eligible articles and full-text review were scrutinised by one investigator (JC). Of those, 271 were ineligible based on criteria and 46 articles were excluded as duplicates, with 21 articles included in this review.

**Figure 1.**
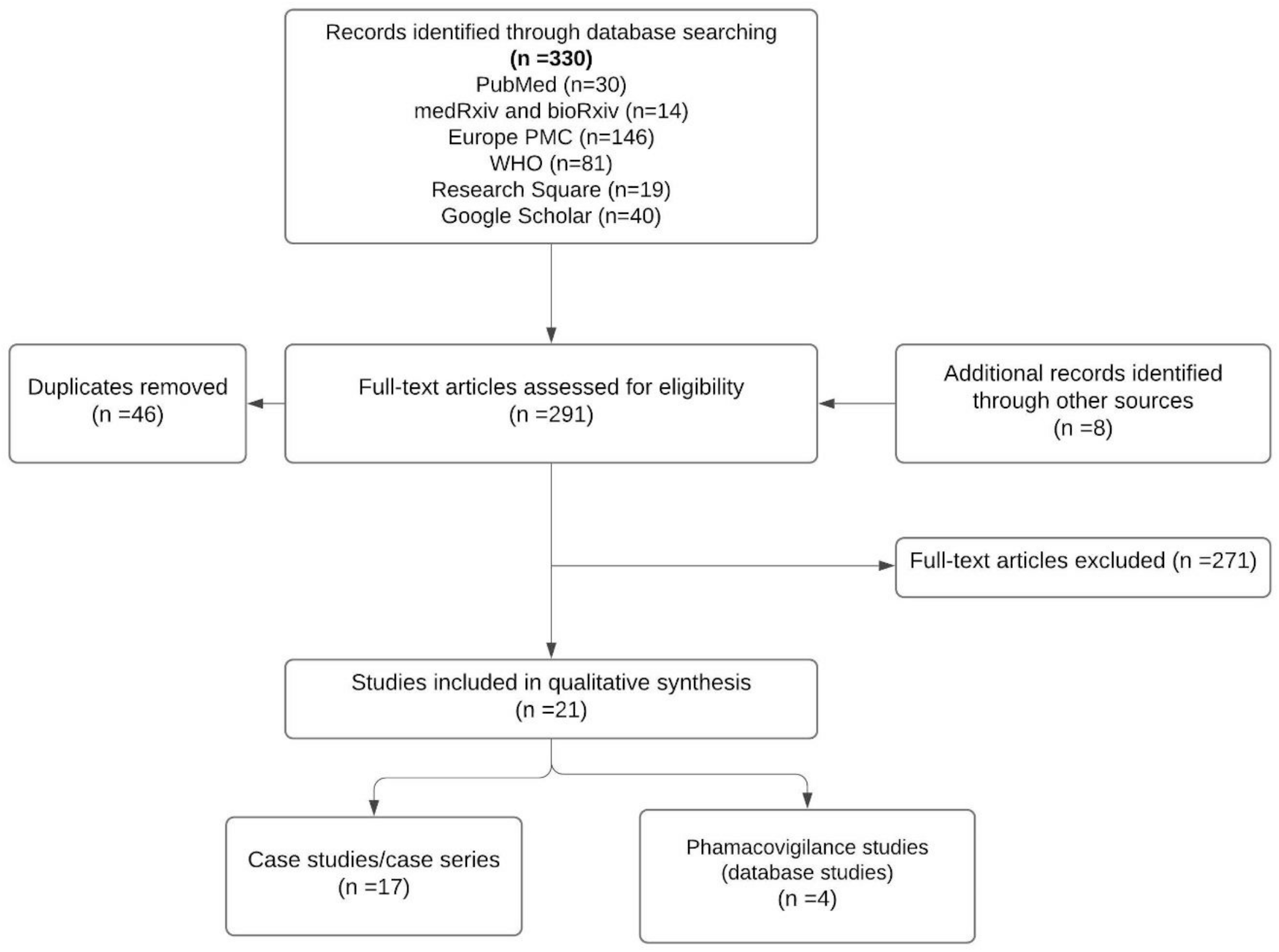
PRISMA search flowchart

### Data extraction

The following information was extracted from included studies: author and year of publication, type of study (cohort, case report or case series), number of cases, case source, patients’ sex, median age and range, vaccine type, percentage of patients presenting symptoms after second vaccination, percentage of patients with symptoms resolved, median and range hospitalisation days and diagnoses of cardiac adverse effects. For the diagnostic criteria and treatment, we extracted the patients’ symptoms, troponin and c-reactive protein blood serum concentration, and observations from electrocardiogram, echocardiogram and cardiovascular magnetic resonance, as well as treatment provided. These data were extracted to a spreadsheet by one reviewer (JC) and checked by two reviewers (JW, AD). They reached agreement regarding any conflicting results.

### Quality evaluation

Quality evaluation of case reports/case series was assessed using the case-report (CARE) 13-item guideline ^22^. This guideline comprises 30 sub-items. Each item ratings are ‘yes’ or ‘no’, and the final quality evaluation score was the sum of sub-items. The quality evaluation was performed independently by two reviewers (JC and AD) and in case of disagreement, a third reviewer (LF) made the final decision. Quality evaluation for each case report/case series are available in supplementary material (Table S2).

### Data synthesis

We considered data from case reports/case series separately from the data extracted from databases reporting adverse effects of vaccination against SARS-CoV-2. Data from three passive surveillance databases were used in included studies; the European Medicines Agency’s European database of suspected adverse drug reaction reports (EudraVigilance, covering European Union and European Economic Area countries)^21^, the Centers for Disease Control (CDC) Vaccine Adverse Event Reporting System (VAERS, covering the USA), ^20^ and the World Health Organisation’s global Individual Case Safety Report Database, VigiBase ^23^. More than 130 countries contribute to VigiBase although more than 85% of adverse effects caused by BNT162b2 (Pfizer) vaccine were reported from countries in Europe and America ^24^. As above, we only included the most recent studies from these databases. Data were available from one active surveillance system, the CDC’s Vaccine Safety Datalink (VSD)^25^ which is a collaboration between the CDC and eight managed care organisations covering 3.6% of the US population. Participating VSD sites file weekly reports of medically-attended (clinic, emergency department or hospital) adverse events of COVID-19 vaccination for total 10.2 million participants aged ≥12 years.

#### Additional data

We additionally updated estimates from EudraVigilance ^21^ and VAERS ^20^. We calculated crude reporting rates for cardiac adverse events per million doses administrated used the denominator being the sum of first dose and all fully vaccinated CYP as of 6 November 2021 for European Union and European Economic Area countries ^16^ and 31 October 2021 for the USA.^15^ For the USA, but not EU countries, we were also able to stratify adverse effects and calculate crude rates by sex and by dose, using data on proportional US adolescent vaccine uptake by sex following previous reports.^26^ For EU/EEA countries, we considered a time lag of 7-days to consider both reporting lag and a pre-defined 7-day risk window, following previous reports^14^; for USA VAERS, we used the vaccination month reported. For this dataset, we were able to repeat these analyses by month of vaccination from December 2020 to October 2021.

## Results

Our review included 21 articles ^14 23 27-44^ together with updated data from EudraVigilance ^21^ and VAERS ^20^.

### Case series/case reports

Seventeen articles were case reports or series that described between one ^31 34 35 40^ and 31 ^32^ patients aged <18 years, with a total sample of 127 (see Table 1). Most were from the USA (82%) ^28-30 32-38 40-43^, with 2 from Israel ^27^ ^39^ and 1 from Germany ^31^. 92.1% of cases were male and the median age ranged between 15 and 17 years. Sixteen described occurrence of cardiac adverse events after vaccination with BNT162b2 (Pfizer) vaccine and one study included cases after Pfizer or Moderna (mRNA-1273) vaccines ^32^. Events were observed after administration of second dose (92.1%) with onset symptoms within 2 ^27 28 32 33 39 41-43^ and 4.5 ^30^ days (median).

**Table 1.**
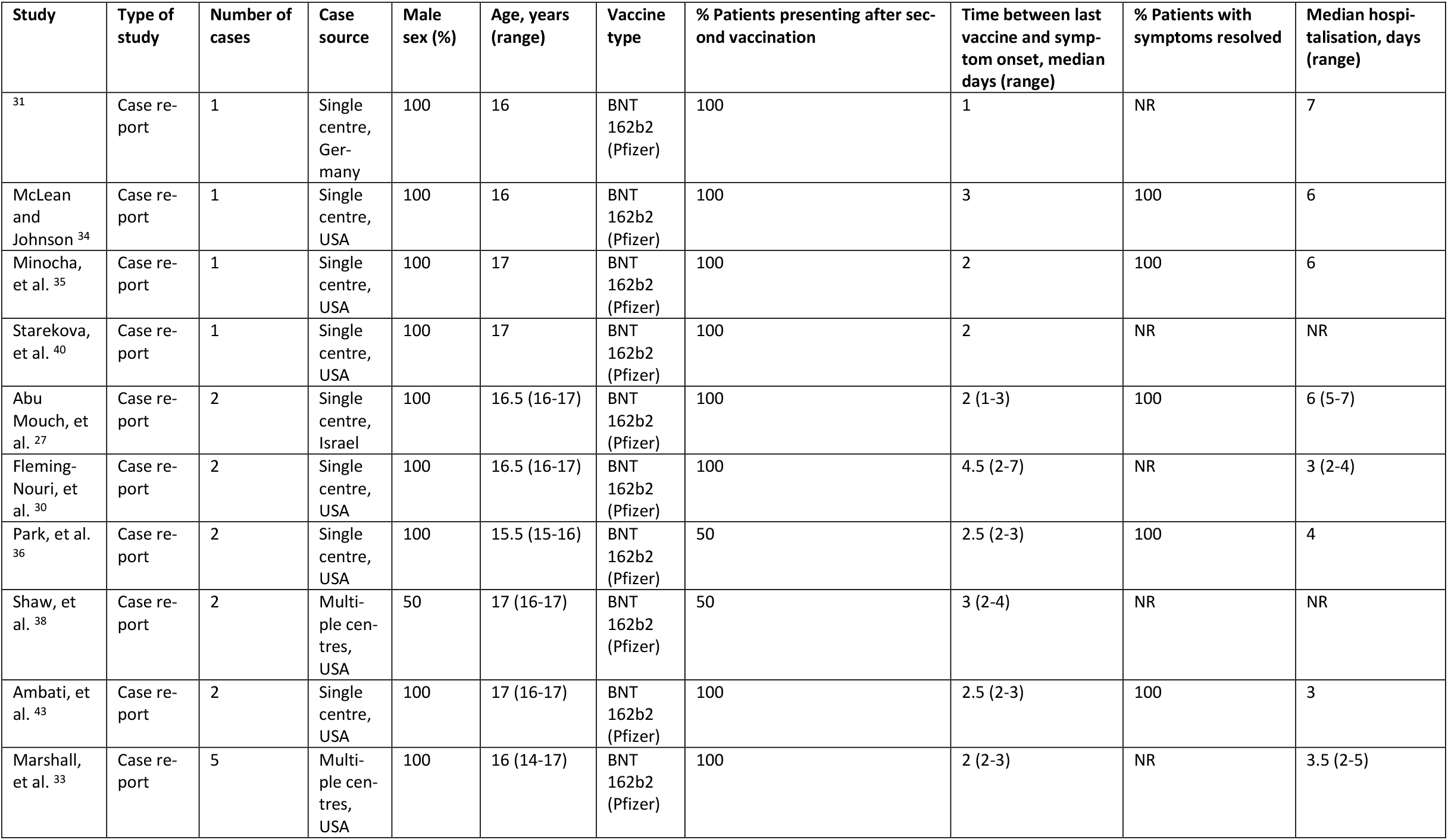
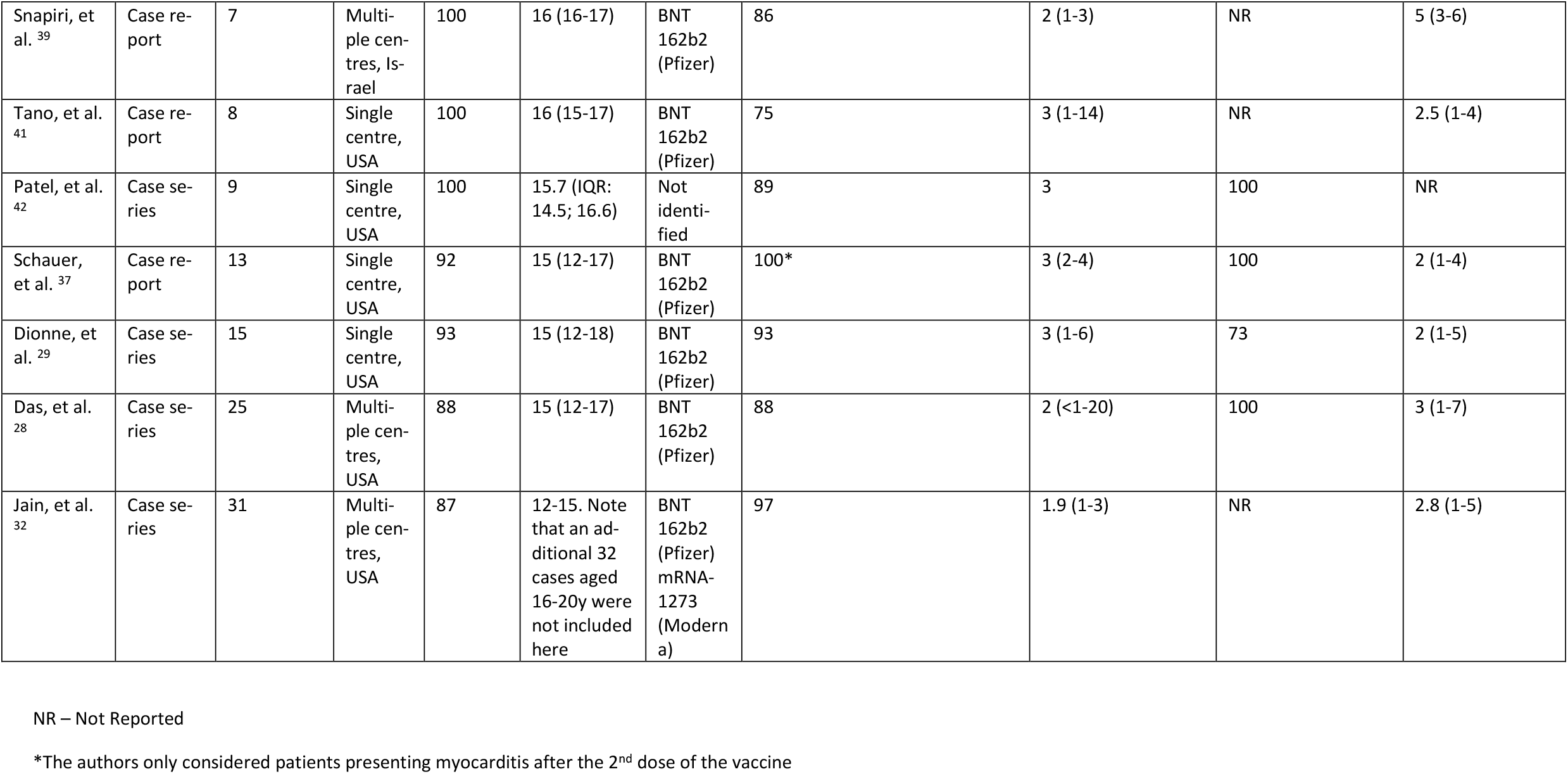
Characteristics and outcomes of the studies describing patients with cardiac adverse effects (myocarditis, pericarditis or myopericarditis related to COVID-19 vaccination.

Sixty-four patients had myocarditis ^27 29 31-33 35 36 38 40 42 43^ and 56 myopericarditis.^28 30 34 37 39 41^ A further 14 patients aged 12-20 years in one study^32^ were considered to have clinically suspected myocarditis although the number 12-17 years was not specified. No patients 0-17 years died of cardiac adverse effects after vaccination in any study. Where reported, the median hospitalisation days ranged between 2 ^29 37 41^ and 7 ^31^ days, and all made full clinical recovery ^28 30-37 39 42 43^ except 14 patients ^29 33 42^ In the latter cases, at the time of discharge, three patients had mildly elevated troponin levels, one had persistent borderline low left ventricular systolic function on echocardiogram (EF 54%) and another had non-sustained ventricular tachycardia. ^29^ Three patients had abnormal cardiovascular magnetic resonance (CMR) results with late gadolinium enhancement (LGE). ^29^ Six patients’ ECGs, at follow-up, revealed persistent changes, either nonspecific T-wave changes ^33^ ^29^, T-wave inversion ^29^ or a repolarisation abnormality. ^42^ Clinical and laboratory findings and treatment given are summarized across the 17 case studies/series in Table 2. Presentation was most commonly with acute chest pain followed by fever and shortness of breath (Table 2). All patients had an elevated troponin (more significant in those with myocarditis) and most a raised CRP. 73.8% of patients had an abnormal ECG at presentation, typically with ST elevation or diffuse ST changes in addition to T wave inversion. Echocardiographic assessment showed normal left ventricular wall geometry and preserved systolic function in the majority. CMR imaging was undertaken in 91 cases with findings suggesting myocardial injury including 56 (61.5%) with myocardial oedema and 82 (90.1%) with LGE. Systolic or diastolic dysfunction was only evident in 5.6% and 7.7%, by echo or CMR assessment, respectively, and 11.3% and 20.9% of patients were noted to have pericardial effusion also by echo or CMR assessment.

**Table 2.**
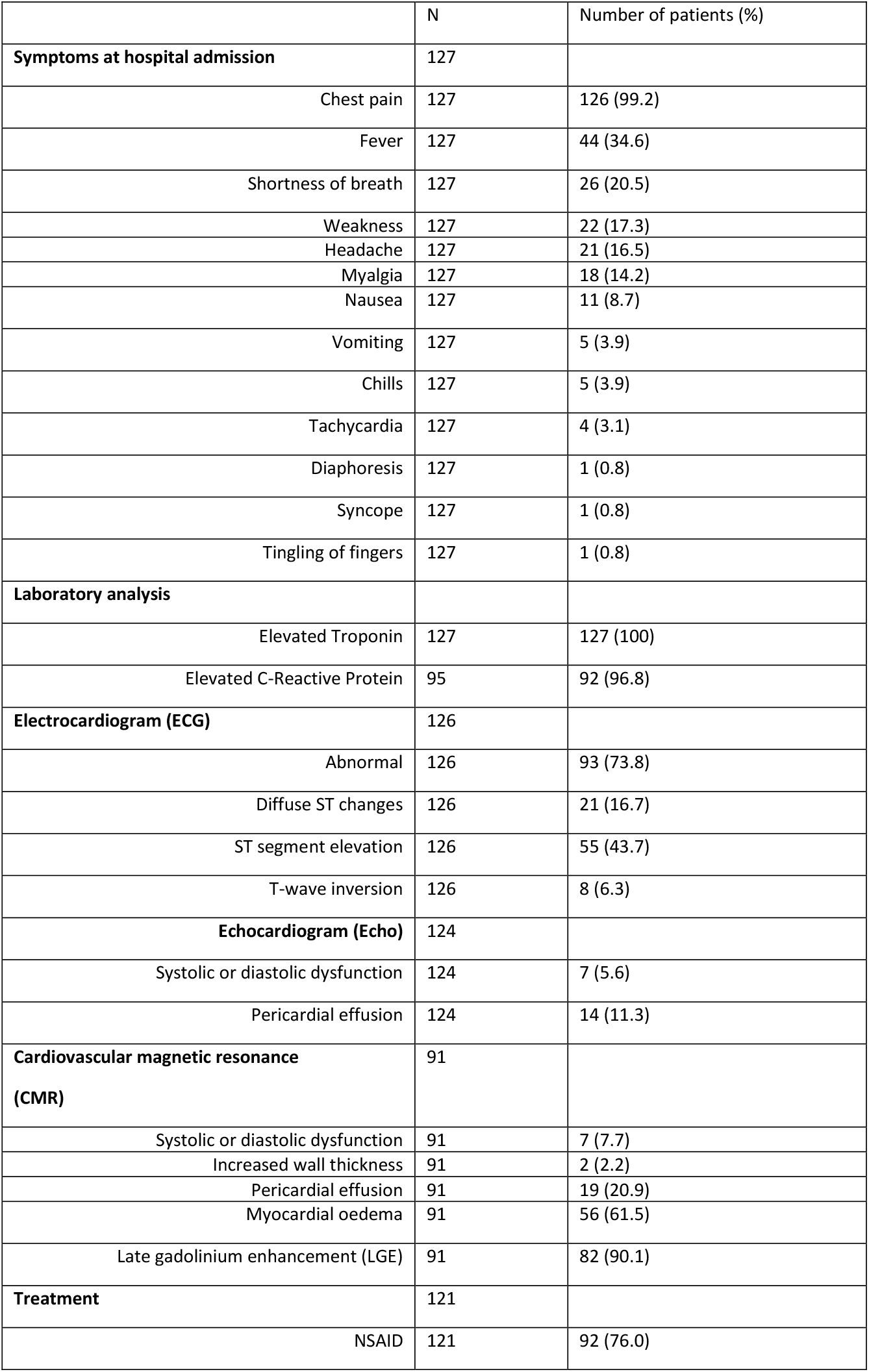
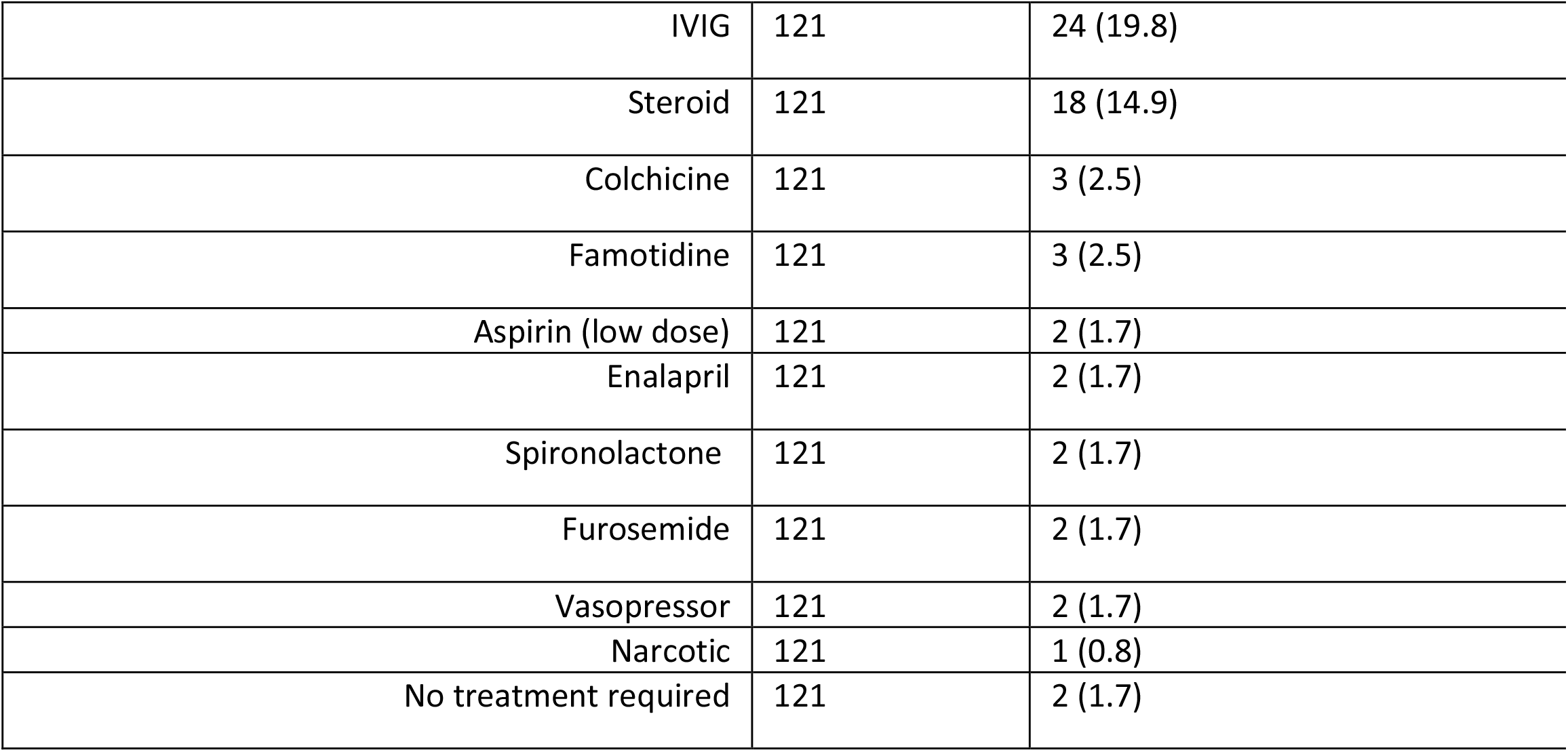
Summary of the most common symptoms presented by patients diagnosed with cardiac adverse effects after being vaccinated against SARS-CoV-2

Treatment most commonly used nonsteroidal anti-inflammatory agents (NSAIDs; 76.0%). Small numbers (14.9%) were treated with corticosteroids. Immunomodulatory therapy in the form of IVIG was used in 19.8% as treatment for myocarditis. Small numbers received diuretics or angiotensin-converting enzyme (ACE) inhibitors (Enalapril; 1.7%) to treat left ventricular systolic dysfunction, or colchicine (2.5%) to prevent recurrent pericarditis. No patients required inotropic or mechanical support. Two (1.7%) patients required no treatment.

### SARS-CoV-2 vaccination adverse effects reported from databases – systematic review

Three included studies used data extracted from passive vaccine surveillance systems, the EudraVigilance^44^, VAERS^20^ ^44^ and VigiBase ^23^ databases with most recent data being from 30 June 2021 (VigiBase ^23^), 6 August 2021 (EudraVigilance ^44^) and 18 August 2021 (VAERS ^20^). We included also a study using Vaccine Safety Database (VSD) data, which included three reports. ^45-47^

VAERS data on reported cases of myocarditis per million amongst US 12-15 year old boys were 4.8 and 42.6 for first and second dose of the Pfizer vaccine^14^. For girls of the same age group, the cases of myocarditis were 0.5 and 4.3 per million of per million of first and second dose of Pfizer vaccine, respectively ^14^. For age group 16-17, the number of cases increased. After the first dose, there were 5.2 and 0.0 cases of myocarditis per million of Pfizer vaccine administrated, in boys and girls respectively ^14^. After the second dose, the numbers increased to 71.5 and 8.1 cases per million of Pfizer vaccine administrated, respectively ^14^. Cases of myocarditis after administration of Moderna vaccine were only reported in boys aged 16-17; after the second dose, 31.2 cases per million of Moderna vaccine administrated.^14^

Only one study published data extracted from EudraVigilance, reporting 61 cases of myocarditis and 14 of pericarditis, in CYP aged 12-17 years, after at least one dose of Pfizer or Moderna vaccines, by 6 August 2021 ^44^. These adverse cardiac effects affected mostly boys (myocarditis: 87%; pericarditis: 64.3%) and were predominantly after Pfizer vaccine (myocarditis: 98.3%; pericarditis: 100%) rather than Moderna vaccines ^44^. Relationship to dose-number and frequency of events per million doses were not available for 12-17 year olds. Whilst some deaths were reported post-vaccine across all ages, data were not available to identify whether within the CYP age-range.

The study from VigiBase reported 291 events after the Pfizer vaccine and 3 after the Moderna vaccine; 249 events were myocarditis and 45 were pericarditis, with no deaths amongst the 12-17 year age-group^23^. Compared with females, the odds of myocarditis was 12.0 (95% CI: 7.9-18.3) for males 12-17 years, with odds for pericarditis amongst boys compared with girls being 12.7 (95% CI: 4.5-35.5) ^23^.

The study using VSD data (3 reports)^45-47^ identified 22 events in the 21 days after SARS-CoV-2 vaccination amongst US 12-17 year olds in data from December 2020 to 21 August 2021. These were a subgroup of a larger sample of 100 cases amongst 12-39 year olds of which 78 (78%) were considered to be true cases of myo- or pericarditis after expert review. Amongst the 22 proven cases in 12-17 year olds, 50% (n=11) had acute myocarditis, 45% (10) myopericarditis and 5% (1) had acute pericarditis. All (22) presented with chest pain and 64% (14) had shortness of breath. 95% (21) had elevated troponin, 73% (16) had an abnormal ECG and 43% (9/21) had an abnormal echocardiogram. 82%(18) were male with a median onset of symptoms of 2 days. 86% (19) were hospitalised (32% (7) to intensive care) and all were discharged home, with 59% discharged within 2 days.^45^ Using additional data to 10 September 2021^46^ on 12-17 year olds, they estimated that for the Pfizer vaccine only (30 confirmed events), there was a significant excess of 29.6 events per million vaccine doses across both sexes and doses, when compared with vaccinated adolescents 22-42 days after each dose, adjusted for site, sex, ethnicity and date. Estimates were 3.8 per million for first doses (not significantly different to zero) and 56.7 per million (significantly different to zero, confidence intervals not reported) for second doses. Three-month follow-up data were available on 16 of the cohort after Pfizer-BioNTech vaccination, of whom 4 (25%) were still symptomatic and 2 (13%) remained on medication, although no data were available on a control group.

### Updated data on SARS-CoV-2 vaccination adverse effects from VAERS and EudraVigilance

We updated the data on cardiac events post vaccination amongst 0-17 year olds in VAERS and EudraVigilance to the latest available data at the time of writing (17 November 2021) (Table 3; Figure 2).

**Table 3.**
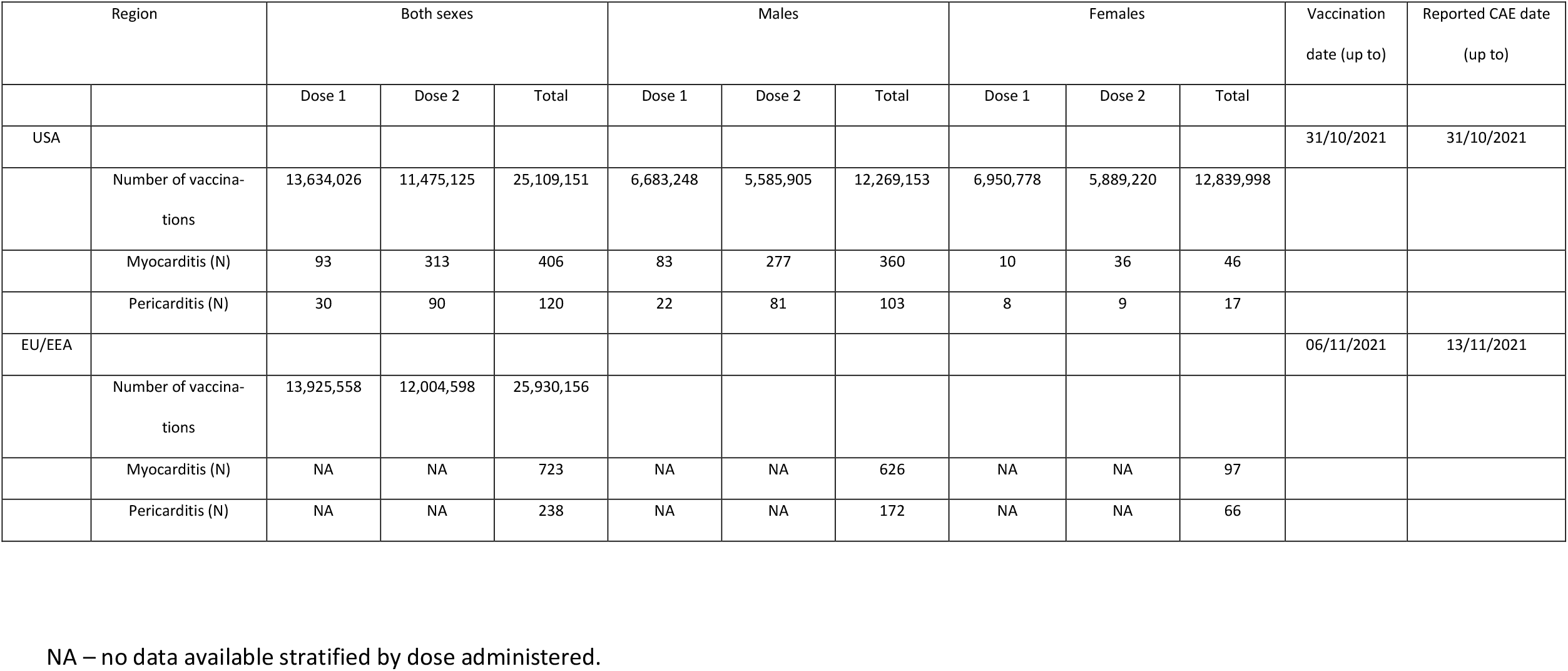
Updated data on SARS-CoV-2 vaccination cardiac adverse effects (CAE) (myocarditis and pericarditis) from VAERS and EudraVigilance for children and young people aged between 6 and 17 years old (VAERS) and 3 and 17 years old (EudraVigilance).

| Region |  | Both sexes |  |  | Males |  |  | Females |  |  | Vaccination<br>date (up to) | Reported CAE date<br>(up to) |
| --- | --- | --- | --- | --- | --- | --- | --- | --- | --- | --- | --- | --- |
|  |  | Dose 1 | Dose 2 | Total | Dose 1 | Dose 2 | Total | Dose 1 | Dose 2 | Total |  |  |
| USA |  |  |  |  |  |  |  |  |  |  | 31/10/2021 | 31/10/2021 |
|  | Number of vaccinations | 13,634,026 | 11,475,125 | 25,109,151 | 6,683,248 | 5,585,905 | 12,269,153 | 6,950,778 | 5,889,220 | 12,839,998 |  |  |
|  | Myocarditis (N) | 93 | 313 | 406 | 83 | 277 | 360 | 10 | 36 | 46 |  |  |
|  | Pericarditis (N) | 30 | 90 | 120 | 22 | 81 | 103 | 8 | 9 | 17 |  |  |
| EU/EEA |  |  |  |  |  |  |  |  |  |  | 06/11/2021 | 13/11/2021 |
|  | Number of vaccinations | 13,925,558 | 12,004,598 | 25,930,156 |  |  |  |  |  |  |  |  |
|  | Myocarditis (N) | NA | NA | 723 | NA | NA | 626 | NA | NA | 97 |  |  |
|  | Pericarditis (N) | NA | NA | 238 | NA | NA | 172 | NA | NA | 66 |  |  |

NA – no data available stratified by dose administered.

**Figure 2.**
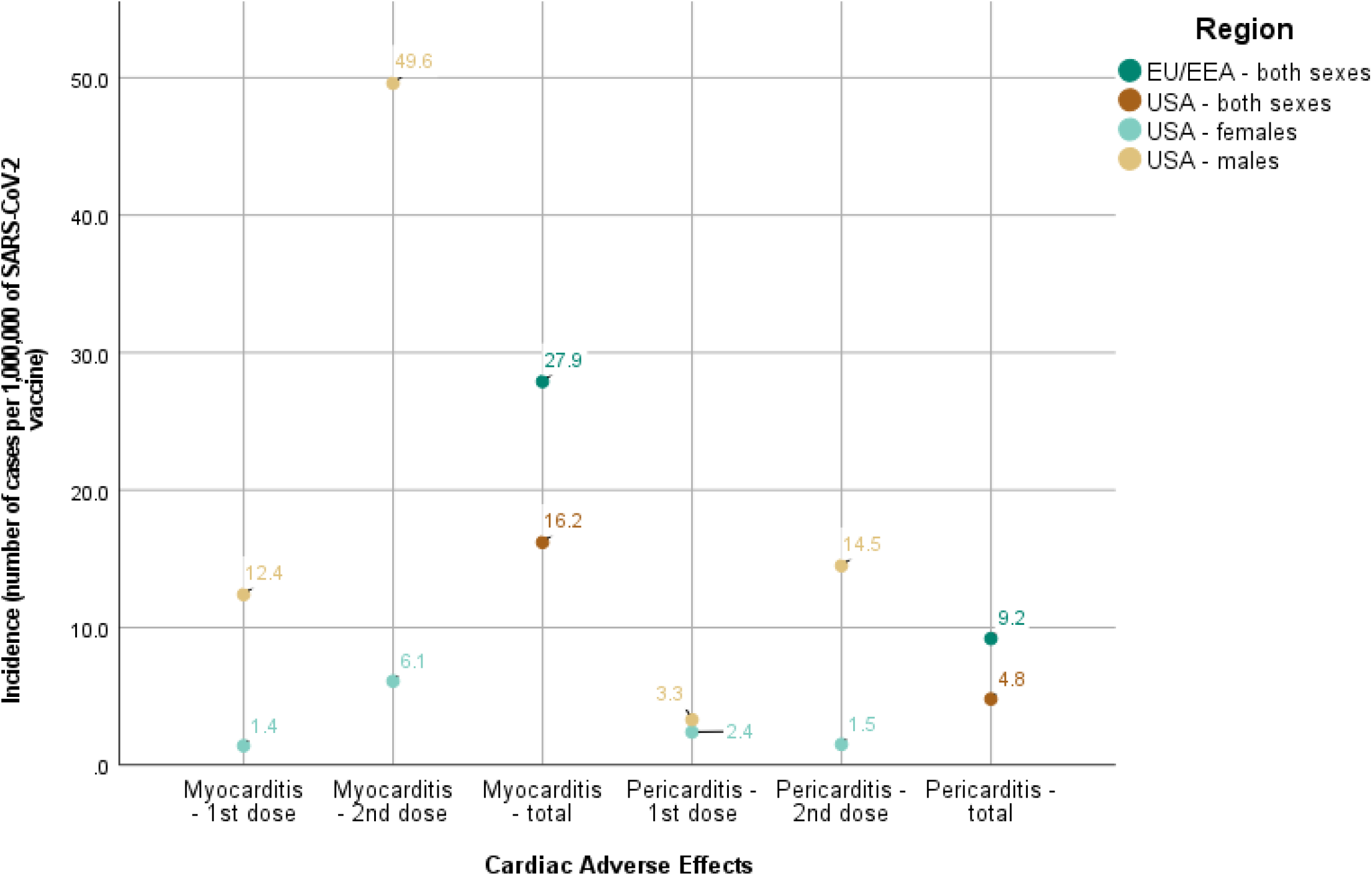
Number of cases of myocarditis and pericarditis reported per 1,000,000 of SARS-CoV-2 vaccines administered to children and adolescents (≤17 years old) reported to EudraVigilance (EU/EEA) and Vaccine Adverse Event Reporting System (VAERS) (USA).

In the USA, 406 cases of myocarditis and 120 of pericarditis were reported to VAERS ^20^ by 31 October 2021. In the EU/EEA, 723 cases of myocarditis and 238 of pericarditis were reported by 13 November 2021 (Table 3). Total number of cardiac events were not calculated due to potential duplicate reporting across these categories.

Two reports recorded death after Pfizer vaccination. One from VAERS was a 15 year old male who died of drowning five days after his second vaccine dose. Autopsy identified increased left ventricular wall thickness and a small focus of myocardial inflammation but a normal pericardium. Genetic analysis of 92 genes associated with inherited cardiomyopathies and arrhythmias identified two variants of unknown significance. The second, from EudraVigilance, was a health professional report of the death of a 12-17 year old male after his second vaccine dose, with no further data beyond myocarditis, headache and fever.

Figure 2 shows the reporting rate of myocarditis and pericarditis in these two datasets. The rate in the reporting EU/EEA countries was 27.9 and 9.2 cases respectively of myocarditis and pericarditis per million vaccines administered.^21^ For the USA, the reporting rate was 16.2 and 4.8 cases per million of SARS-CoV-2 respectively.^15^ A higher percentage of male adolescents than females reported cardiac adverse effects for both conditions studied (myocarditis: 86.6% (EU/EEA) ^21^ and 88.7% (USA) ^20^; pericarditis: 72.3% (EU/EEA) ^21^ and 85.8% (USA) ^20^) (Table 3).

Only the USA VAERS provided data on the relationship of adverse events to vaccine dose; the rate of myocarditis per million after first dose was 12.4 for boys and 1.4 for girls, and 49.6 for boys and 6.1 for girls after the second dose. The reporting rate of pericarditis followed a similar trend. Across both sexes, the majority of cardiac events were reported after the second dose (myocarditis: 77.1%; pericarditis: 75.0%).

Again, only VAERS provided data on time to onset of symptoms and duration of hospitalisation. Median onset of symptoms was two days (range: 0 to > 120 days) after immunisation ^20^. The median days that CYP were hospitalised was two (ranging from no need of hospitalisation to two months) ^20^.

VAERS data for myocarditis and pericarditis events by month of vaccination are shown in Figure 3. For myocarditis, these show a trend for events after first and second doses to be higher soon after initiation of that element of the vaccine schedule (February 2021 for first dose, April 2021 for second doses). Incidence of events after second dose fell each month from July onwards, a pattern also seen in females. In the most recent months (September-October 2021), incidence of myocarditis after first doses was 7.6-12.3 in males and 0 in females, and after second dose 13.1-31.1 for males and 1.3-2.5 per million for females.

**Figure 3.**
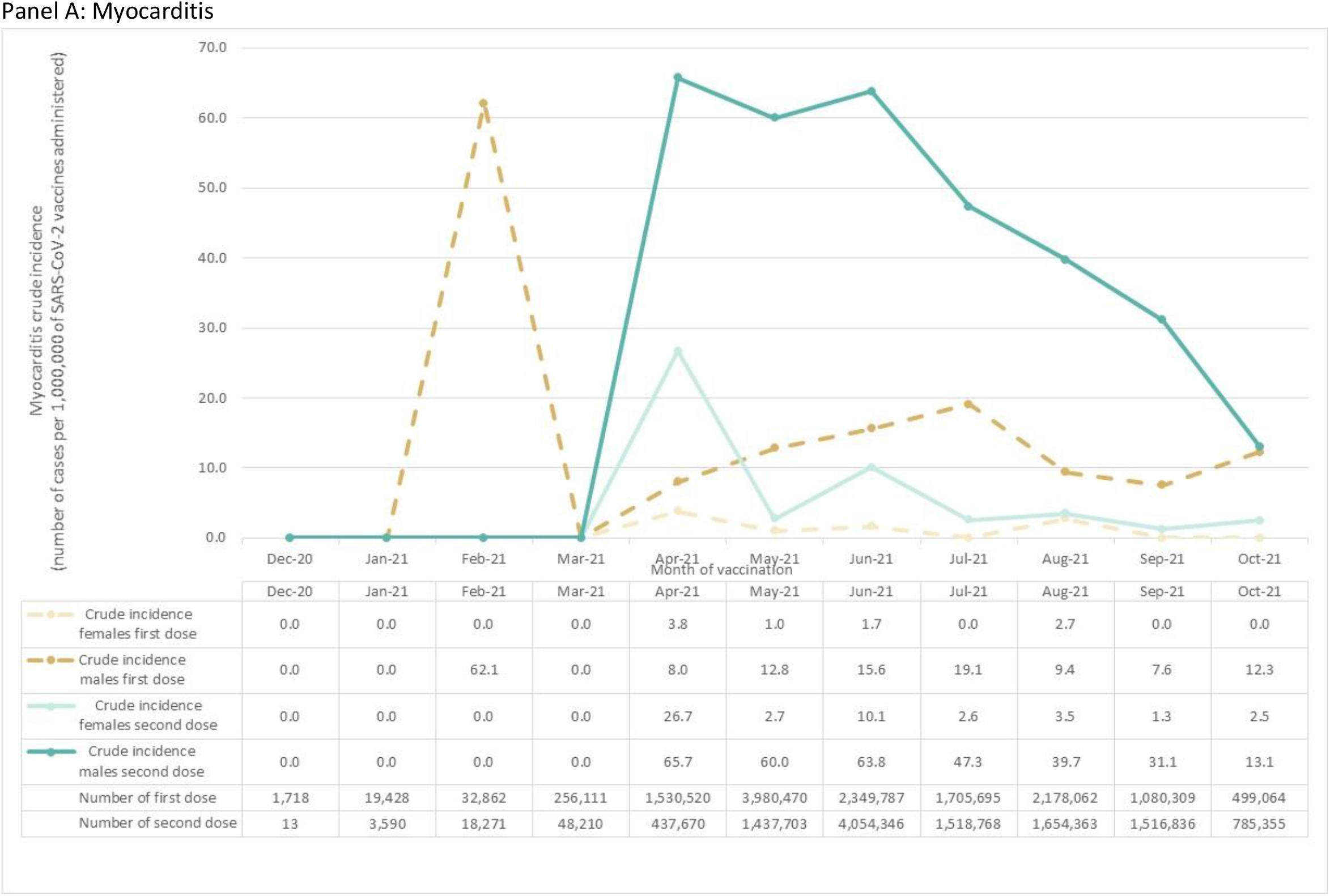
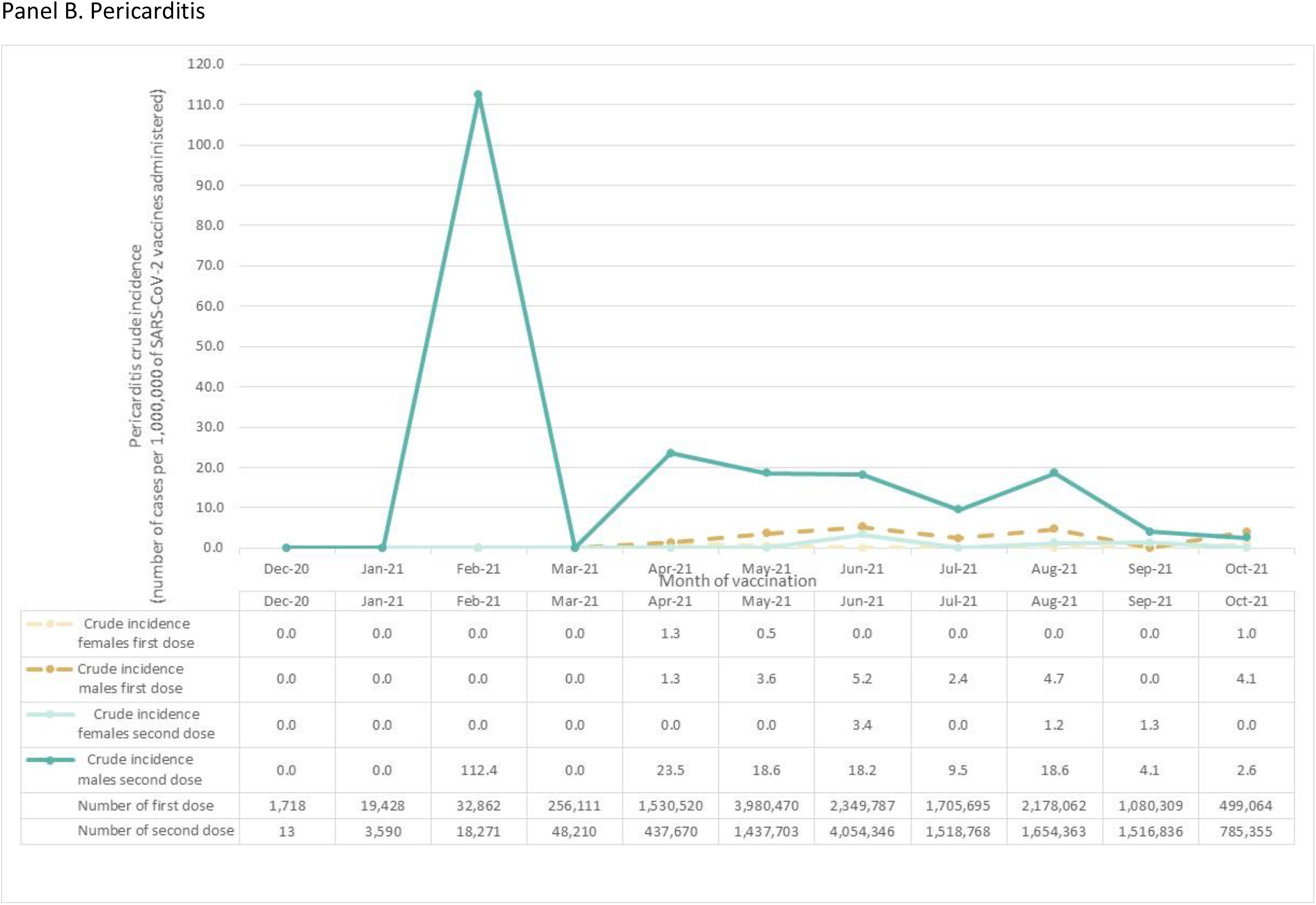
Crude incidence of cases of myocarditis and pericarditis reported to the Vaccine Adverse Event Reporting System (VAERS) per million vaccines, by month of vaccination

Incidence of pericarditis after the second dose in males appeared to follow a similar trend, being highest in February and April 2021 and falling thereafter. Incidence in the most recent two months was 2.6-4.1 per million for males and 0-1.3 for females.

## Discussion

Our systematic review provides a comprehensive and up-to-date (November 2021) synthesis of reported data on cardiac adverse effects in CYP < 18 years after SARS-CoV-2 immunisation. Detailed clinical data were available on 127 cases across 17 studies, active surveillance data on 30 confirmed cases, and passive surveillance data on 1129 reports of myocarditis and 358 reports of pericarditis from across the USA and EU/EEA. It is highly likely that some or all of the clinical cases and active surveillance cases were contained within the passive surveillance reports. Events were also reported through the WHO VigiBase database, although the degree to which these events overlapped with the US and EU/EEA reports is unclear as these countries make up around 70% of VigiBase reports.

Clinical data were highly consistent across the three sources of data. The majority of cases occurred in males aged 15-17 years old who presented with chest pain, elevated serum troponin and C-reactive protein and an abnormal ECG with ST elevation. Fewer presented under age 15 years. We identified no cases under 12 years of age although very few doses have been given in this age group. Most presented with chest pain around 2 days after vaccination, predominantly their second dose. Almost all had high troponin levels, 73.8% had an abnormal ECG with variable echocardiography findings. Approximately 85% were hospitalised for short periods, with the great majority discharged within 2-7 days without treatment other than non-steroidal anti-inflammatory drugs. While longer-term data were only available on 16 young people from the VSD study, few remained on medication at 3 months. The clinical picture of myocarditis post-COVID-19 vaccination amongst 12-17 year olds was broadly similar to that previously reported across all ages.^11^ ^12^

There were no deaths in the clinical series nor in the active surveillance series, and none in the clinical series required significant cardiovascular or respiratory support. There were two male deaths reported in the passive surveillance data, although a causal link cannot be attributed to vaccination, given that all deaths within a 21 day period after vaccination maybe reported regardless of causation. It is important to note that in 2019 in the USA there were approximately 12 deaths males aged 12-17 years each day, with approximately 250 occurring in the average 21 day period, from a range of traumatic and medical causes.^48^ Given that over 40% of 12-17 year olds in the USA have had at least one dose, ^26^ this suggests that there are likely to be a considerable number of deaths of adolescent boys soon after vaccination for reasons entirely unconnected with vaccination. Broader analysis of VAERS reports of deaths from myocarditis associated with COVID-19 vaccination across a broader age-group is reassuring that almost all reported cases are not vaccination-related. As of early November, 8 deaths were identified in those aged <30 years with possible concern for myocarditis after approximately 86 million vaccine doses; 3 were assessed as not being myocarditis, 3 as due to infectious (i.e. non-vaccine) myocarditis whilst evaluation was ongoing in 2 cases.^47^

Data from active surveillance suggest a significant excess of confirmed myo- or pericarditis events in the 21 days post-vaccination compared with matched adolescents 22-42 days after vaccination, supporting a causal link for myocarditis with the Pfizer-BioNTech vaccine although numbers of events were low in this age-group. Passive surveillance data from VAERS and EudraVigilance provide an estimate of the reporting rate for unconfirmed events and cannot infer causality. These differences in data quality and estimation methods means that estimates are not directly comparable across the datasets, although estimates were broadly similar. Overall estimates across both sexes and both doses across the period were approximately 30 per million for myo-pericarditis for VSD and 17 and 28 per million for myocarditis in VAERS and EudraVigilance respectively. Estimates for events post first dose were approximately 4 per million from VSD and 1.4 and 12.4 for females and males respectively in VAERS.

However the time trends we identified in VAERS data for males suggest that the summary estimates across the study period are highly likely to be biased upwards. Estimates were higher amongst males after the second dose across the study period, however the monthly crude estimate decreased notably in the most recent 3 months. Whilst the most recent month may be lower due to incomplete reporting, previous studies showing higher levels of reporting in VAERS following publicity relating to claimed adverse events,^49^ suggest that lower estimates from August and September 2021 (31.1-39.7 per million second doses) may be less biased, although trends may be continuing. It is unclear whether similar trends have occurred in notifications to EudraVigilance as trend data are not publicly available. Whilst there was no similar trend for events post first dose, it is notable that passive surveillance reports from the UK, which began vaccinating healthy adolescents from September 2021, more recently than the USA or many EU countries, identified a reporting rate of 0.8 events per million amongst 0-17 year olds after approximately 2.8 million doses of the Pfizer-BioNTech vaccine, the great majority being first doses.^50^

Post-vaccination myocarditis must be seen in the context of myocarditis related to SARS-CoV-2 infection, post-COVID syndromes and other causes of myocarditis. A large Israeli study found the risk of vaccine-related myocarditis was found to be approximately 4-fold less than the risk of myocarditis due to SARS-CoV-2 infection across age-groups,^9^ although similar data are not available for the adolescent age-range. The post-COVID-19 Paediatric Inflammatory Multisystem Syndrome (PIMS) has well described cardiovascular involvement. Paediatric patients may present with cardiogenic shock, ECG abnormalities, myocardial dysfunction and coronary artery dilation ^10^, with 71-75% patients being admitted to Paediatric Intensive Care Unit (PICU) ^51^ ^52^ although the risk of death is very low.^53^ Further, the clinical picture and outcomes of myocarditis post-COVID-19 vaccination appear quite different to established causes of acute myocarditis in CYP, which occurs in around 8 per million CYP^54^ and has a higher incidence of left ventricular dysfunction and arrhythmia, median length of stay 6-7 days,^54^ ^55^ and mortality of up to 3-8%.^54 56 57^

Other vaccines have rarely been linked with cardiac adverse effects.^58 59 60^ A reviewed of VAERS reports of myopericarditis after vaccination between 1990 and 2018 found 99 reports of myopericarditis in CYP, with most cases reported after *Haemophilus influenza* type b (22, 22%) and hepatitis B (18, 18%) vaccination ^60^. Most cases reported were in males (55, 56%), of high severity (40, 40%), and resulted in death in 55% of cases,^60^ although this may reflect biased reporting only of severe cases.

### Limitations

Our findings are subject to a number of limitations inherent in systematic reviews and in the data sources used. Publication bias towards greater severity is likely amongst the cases reported in the 17 case study/series studies, which may not be representative of the less severe cases.

Estimates from vaccine passive surveillance systems such as VAERS and EudraVigilance provide a reporting rate rather than an estimate of excess events, and are crude indicators of risks related to vaccines.^61^ They cannot attribute causality between vaccine and reported event, lack a comparison group, data quality is highly variable and they are open to a series of biases from over- and -under-reporting.^62^ VAERS in particular has been criticised as open to over-reporting by those opposed to COVID-19 vaccination.^63^ ^64^ However all reports are regularly scrutinized to remove duplicate and reports identified as fraudulent, ^62^ although the extent and timeliness of this is unclear. Passive surveillance systems are most useful for identifying potential safety signals that require further investigation^62^ through more detailed smaller studies using active systems such as the Vaccine Safety Datalink (VSD). The VSD data included here provide an estimate of excess events, but the VSD can lack power to examine small risks in specific populations^25^ such as the CYP studied here.

Our data focus on 12-17 year olds and should not be regarded as informative for younger children, as very few received COVID-19 vaccination during the period of study here. We were unable to include data from some large all-age studies^12^ as these did not provide data specifically for those <18y. We were unable to further breakdown VAERS data by age within the adolescent range; a CDC report from October 2021 reported that pooled risk for 16-17 year olds was higher after the second dose (69.1 per million) than amongst 12-15 year olds (39.9 per million).^47^ We estimated sex-specific reporting rates from VAERS data using proportional differences in vaccine uptake in US adolescents. We were unable to compare risk of cardiac events after the Pfizer-BioNTech and Moderna vaccines in this age group due to lack of data. We were further unable to comment on risk of myocarditis after third doses due to lack of data as no country has introduced booster doses for this age-group.

## Conclusions

Cardiac adverse effects are very rare after use of mRNA vaccination for COVID-19 in CYP <18 years. The great majority of cases are mild and self-limiting without significant treatment, in contrast to viral myocarditis. No data are yet available on children under 12 years and these are awaited as vaccination of 5 to 11 year olds is rolled-out in some countries. Larger detailed prospective longitudinal studies from active surveillance sources on both children and adolescents are urgently needed to better understand the frequency and natural history of the condition.

## Supporting information

Keyword strings Search terms

Quality evaluation of case reports/case series

## Data Availability

All data produced in the present work are contained in the manuscript

