## Supplementary material for "Systematic review of cardiac adverse effects in children and young people under 18 years of age after SARS-CoV-2 vaccination": Keyword strings Search terms

Table S1 – Keyword strings Search terms

| Search engine and database | SARS-CoV-2 vaccination |
|---|---|
| Pubmed | ((pericarditis[Title/Abstract]) OR (myocarditis[Title/Abstract]) OR ("adverse effects" [Title/Abstract]) OR (cardiac[Title/Abstract])) AND (covid-19[Title/Abstract]) AND (vaccin*[Title/Abstract]) AND ((children[Title/Abstract]) OR (adolescent[Title/Abstract]) OR (paediatric [Title/Abstract])) |
| MedRxive<br><br>And BioRxive | abstract or title "pericarditis vaccine" (match all words)<br><br>OR abstract or title "myocarditis vaccine" (match all words)<br><br>OR abstract or title "cardiac vaccine" (match all words)<br><br>and posted between "01 Dec, 2019 and 14 September, 2021" |
| Europe PMC | (ABSTRACT:"myocarditis" OR ABSTRACT:"pericarditis" OR ABSTRACT:"adverse effects" OR ABSTRACT: "cardiac") AND (ABSTRACT:"covid-19") AND (ABSTRACT:"vaccine" OR ABSTRACT: "vaccination") AND (FIRST_PDATE:[2019-12-01 TO 2021-09-14]) |
| World Health Organisation | ((tw:(myocarditis)) OR (tw:(pericarditis)) OR (tw:(cardiac)) OR (tw:("adverse effects"))) AND (tw:(vaccine)) OR (tw:(vaccination))) AND ((tw:(children)) OR (tw:(adolescent)) OR (tw:( pediatric)) OR (tw:( paediatric))) |
| Research Square | Abstract: "myocarditis" |
| Google scholar | covid-19 and vaccine* and (*myocarditis OR cardiac) |

((tw:(myocarditis)) OR (tw:(pericarditis)) OR (tw:("adverse effects"))) AND (tw:(vaccine)) OR (tw:(vaccination)) AND (tw:(children)) OR (tw:(adolescent)) OR (tw:(pediatric)) OR (tw:(paediatric))
