## Supplementary material for "Systematic review of cardiac adverse effects in children and young people under 18 years of age after SARS-CoV-2 vaccination": Quality evaluation of case reports/case series

| Topic | Item | Checklist item description | Isaak | McClean | Minocha et al | Starekova | Abu Mouch et | Fleming-Nouri | Park | Shaw | Ambati | Marshall | Snapiri et al. | Tano | Patel | Schauer et al. | Dionne et al. ( | Das | Jain et al. (2021) |
|---|---|---|---|---|---|---|---|---|---|---|---|---|---|---|---|---|---|---|---|
| Title | 1 | The diagnosis or intervention of primary focus followed by the words "case report" | 0 | 1 | 0 | 0 | 0 | 0 | 0 | 0 | 0 | 0 | 0 | 0 | 0 | 0 | 1 | 0 | 0 |
| Key Words | 2 | 2 to 5 key words that identify diagnoses or interventions in this case report, including "case repo | 0 | 0 | 0 | 0 | 0 | 0 | 0 | 0 | 0 | 0 | 0 | 0 | 0 | 0 | 0 | 0 | 0 |
| Abstract | 3a | Introduction: What is unique about this case and what does it add to the scientific literature? | 0 | 1 | 0 | 0 | 1 | 0 | 1 | 0 | 1 | 1 | 1 | 1 | 1 | 0 | 1 | 1 | 1 |
| (no referen | 3b | Main symptoms and/or important clinical findings | 0 | 1 | 0 | 0 | 1 | 0 | 1 | 0 | 1 | 1 | 1 | 0 | 1 | 1 | 1 | 1 | 1 |
| | 3c | The main diagnoses, therapeutic interventions, and outcomes | 0 | 0 | 0 | 0 | 1 | 0 | 1 | 0 | 0 | 1 | 1 | 0 | 0 | 0 | 1 | 1 | 1 |
| | 3d | Conclusion-What is the main "take-away" lesson(s) from this case? | 0 | 1 | 0 | 0 | 1 | 0 | 1 | 0 | 1 | 1 | 1 | 0 | 0 | 0 | 1 | 1 | 1 |
| Introductior | 4 | One or two paragraphs summarizing why this case is unique (may include references) | 0 | 1 | 1 | 1 | 1 | 1 | 1 | 1 | 1 | 1 | 1 | 0 | 1 | 1 | 1 | 1 | 1 |
| Patient Info | 5a | De-identified patient specific information | 1 | 1 | 1 | 1 | 1 | 1 | 1 | 1 | 1 | 1 | 1 | 1 | 1 | 1 | 1 | 1 | 1 |
| | 5b | Primary concerns and symptoms of the patient | 1 | 1 | 1 | 1 | 1 | 1 | 1 | 1 | 1 | 1 | 1 | 1 | 0 | 1 | 1 | 1 | 1 |
| | 5c | Medical, family, and psycho-social history including relevant genetic information | 0 | 0 | 1 | 0 | 1 | 0 | 1 | 1 | 1 | 1 | 0 | 1 | 0 | 0 | 0 | 1 | 1 |
| | 5d | Relevant past interventions with outcomes | 0 | 0 | 0 | 0 | 0 | 0 | 0 | 0 | 0 | 0 | 0 | 0 | 0 | 0 | 0 | 0 | 0 |
| Clinical Finc | 6 | Describe significant physical examination (PE) and important clinical findings | 0 | 1 | 1 | 1 | 1 | 1 | 1 | 0 | 1 | 0 | 1 | 0 | 1 | 1 | 1 | 1 | 0 |
| Timeline | 7 | Historical and current information from this episode of care organized as a timeline | 0 | 0 | 0 | 0 | 0 | 0 | 0 | 1 | 0 | 0 | 0 | 0 | 0 | 0 | 0 | 0 | 0 |
| Diagnostic a | 8a | Diagnostic testing (such as PE, laboratory testing, imaging, surveys). | 1 | 1 | 1 | 1 | 1 | 1 | 1 | 1 | 1 | 1 | 1 | 1 | 1 | 1 | 1 | 1 | 1 |
| | 8b | Diagnostic challenges (such as access to testing, financial, or cultural) | 0 | 0 | 0 | 0 | 0 | 0 | 0 | 0 | 0 | 0 | 0 | 0 | 0 | 0 | 0 | 0 | 0 |
| | 8c | Diagnosis (including other diagnoses considered) | 1 | 1 | 1 | 1 | 1 | 1 | 1 | 0 | 1 | 1 | 1 | 1 | 1 | 1 | 1 | 1 | 1 |
| | 8d | Prognosis (such as staging in oncology) where applicable | 0 | 0 | 0 | 0 | 0 | 0 | 0 | 0 | 0 | 0 | 0 | 0 | 0 | 0 | 0 | 0 | 0 |
| Therapeutic | 9a | Types of therapeutic intervention (such as pharmacologic, surgical, preventive, self-care) | 0 | 1 | 1 | 0 | 1 | 1 | 1 | 0 | 1 | 1 | 1 | 1 | 1 | 1 | 1 | 1 | 1 |
| | 9b | Administration of therapeutic intervention (such as dosage, strength, duration) | 0 | 0 | 0 | 0 | 0 | 0 | 0 | 0 | 0 | 1 | 0 | 0 | 0 | 0 | 1 | 0 | 0 |
| | 9c | Changes in therapeutic intervention (with rationale) | 0 | 0 | 0 | 0 | 0 | 0 | 0 | 0 | 0 | 0 | 0 | 0 | 0 | 0 | 0 | 0 | 0 |
| Follow-up a | 10a | Clinician and patient-assessed outcomes (if available) | 0 | 0 | 1 | 0 | 1 | 1 | 1 | 0 | 0 | 1 | 1 | 0 | 1 | 1 | 1 | 1 | 1 |
| | 10b | Important follow-up diagnostic and other test results | 0 | 0 | 1 | 0 | 0 | 0 | 0 | 0 | 0 | 1 | 0 | 0 | 1 | 0 | 1 | 0 | 1 |
| | 10c | Intervention adherence and tolerability (How was this assessed?) | 0 | 0 | 0 | 0 | 0 | 0 | 0 | 0 | 0 | 0 | 0 | 0 | 0 | 0 | 0 | 0 | 0 |
| | 10d | Adverse and unanticipated events | 0 | 0 | 0 | 0 | 0 | 0 | 0 | 0 | 0 | 0 | 0 | 0 | 0 | 0 | 0 | 0 | 0 |
| Discussion | 11a | A scientific discussion of the strengths AND limitations associated with this case report | 0 | 1 | 0 | 1 | 0 | 0 | 0 | 0 | 1 | 1 | 1 | 0 | 1 | 0 | 1 | 1 | 1 |
| | 11b | Discussion of the relevant medical literature with references | 1 | 1 | 1 | 1 | 1 | 0 | 1 | 0 | 1 | 1 | 1 | 1 | 1 | 1 | 1 | 1 | 1 |
| | 11c | The scientific rationale for any conclusions (including assessment of possible causes) | 0 | 1 | 1 | 0 | 1 | 0 | 1 | 0 | 1 | 1 | 1 | 1 | 1 | 1 | 1 | 1 | 1 |
| | 11d | The primary "take-away" lessons of this case report (without references) in a one paragraph cor | 0 | 1 | 1 | 1 | 1 | 0 | 0 | 1 | 1 | 1 | 1 | 1 | 1 | 1 | 1 | 0 | 1 |
| Patient Pers | 12 | The patient should share their perspective in one to two paragraphs on the treatment(s) they re | 0 | 0 | 0 | 0 | 0 | 0 | 0 | 0 | 1 | 0 | 0 | 0 | 0 | 0 | 0 | 0 | 0 |
| Informed C | 13 | Did the patient give informed consent? Please provide if requested . . . . . . . . . . . . . . . . . . . . . . . | 0 | 0 | 0 | 0 | 0 | 0 | 0 | 0 | 0 | 0 | 0 | 0 | 0 | 0 | 0 | 0 | 0 |
| TOTAL | | | 5 | 15 | 13 | 9 | 16 | 8 | 15 | 7 | 16 | 18 | 16 | 10 | 14 | 12 | 19 | 16 | 17 |
